# Resuming professional football (soccer) during the COVID-19 pandemic in a country with high infection rates: A prospective cohort study

**DOI:** 10.1101/2020.11.17.20233023

**Authors:** Yorck Olaf Schumacher, Montassar Tabben, Khalid Hassoun, Asmaa Al Marwani, Ibrahim Al Hussain, Peter Coyle, Ahmed Khellil Abbassi, Hani Taleb Ballan, Abdulaziz Jaham Al Kuwari, Karim Chamari, Roald Bahr

## Abstract

**Objectives:** The risk of viral transmission associated with contact sports such as football (soccer) during the COVID-19 pandemic is unknown. The aim of this study was to describe the infective and immune status of professional football players, team staff and league officials over a truncated football season resumed at the height of the COVID-19 pandemic in a country with high infection rates and to investigate the clinical symptoms related to COVID-19 infection in professional football players.

**Methods:** Prospective cohort study of 1337 football players, staff and officials during a truncated football season (9 weeks) with a tailored infection control program based on preventive measures and regular SARS-CoV-2 PCR swab testing (every 3-5 days) combined with serology testing for immunity (every 4 weeks). Clinical symptoms in positive participants were recorded using a 26-item, Likert-scale-based scoring system.

**Results:** During the study period, 85 subjects returned positive (cycle threshold (cT)≤30) or reactive (30<cT<40) PCR tests, of which 36 were players. The infection rate was consistent with that of the general population during the same time period. More than half of infected subjects were asymptomatic, and the remaining had only mild symptoms with no one requiring hospitalization. Symptom severity was associated with lower cT values. Social contacts and family were the most common sources of infection, and no infection could be traced to training or matches. Of the 36 infected players, 15 presented positive serology during the study period.

**Conclusion:** Football played outdoors involving close contact between athletes represents a limited risk for SARS-CoV-2 infection and severe illness when preventive measures are in place.

## Introduction

The severe acute respiratory syndrome coronavirus 2 (SARS-CoV-2), the agent causing coronavirus disease 2019 (COVID-19), is responsible for a global pandemic. The key preventive intervention imposed by governments world-wide, social distancing, has resulted in severe disruption of most public and private sectors, including sports (1,2).

While training and competitions are gradually resuming around the world, the risks associated with COVID-19 for athletes remain uncertain. The risk of transmission playing contact sports is also unknown. In addition to the lungs, the disease can also affect the cardiovascular system and the blood (3,4). Both organ systems are of crucial importance for sporting performance. Some research suggests that the immune response of athletes might be impaired under heavy training which could hypothetically lead to more severe disease progression (5). On the other hand, professional athletes do not typically possess any of the predisposing factors for severe COVID-19 disease (6). Still, there is limited information on the clinical outcomes for athletes contracting COVID-19.

Data from population studies show that SARS-CoV-2 mainly spreads via respiratory droplets and direct contact and disease severity might be impacted by the viral load taken up at the moment of infection (7). In team sports, personal contact between players is inevitable and a “safe distance” cannot always be maintained. Football players remain at a distance of >1.5 m of another player for the vast majority of the game (8) and physical encounters are brief (tackling and heading duels). Still, the heavy, unprotected breathing during exercise generates more droplets than normal respiration, increasing the risk of exposure (9).

Meyer et al. (10) have recently described the successful reopening of the German professional football league, but in a situation where the risk of viral transmission was low, with only about 5 new cases per 100 000 inhabitants per week in Germany. This study describes the infective and immune status of professional football players, team staff and league officials over a truncated football season resumed at the height of the COVID-19 pandemic in a country with high transmission risk. We also examine the clinical course and symptoms in professional football players afflicted by COVID-19 infection.

## Methods

This prospective cohort study included players, staff and referees of the national professional league (Qatar Stars League, QSL). It was conducted between June 8^th^, 2020 and September 2^nd^, 2020 in Qatar (population: 2.8 million). With few exceptions, players are full-time professionals. Teams are typically supported by 20-25 staff members. The regular season started in August 2019 with 12 teams in Division 1 and 5 teams in Division 2, and planned to be completed by May 2020. Due to the pandemic, the season was interrupted on March 16^th^, 2020.

### Return-to-competition protocol

To allow resumption of the football league, the QSL decided in May 2020 to implement a return-to-competition protocol to complete the season (11). The mainstays of this protocol were strict hygiene measures and regular testing. All players, staff and referees signed written commitments to adhere to these measures; violations were subject to fines from the league.

The protocol included an initial two-week hotel quarantine period, where the teams trained without physical contact. Upon completion and for the rest of the season, players and staff were allowed to live their normal life with a signed pledge to adhere to home quarantine when not training or playing and limit social contacts whenever possible. Other country-wide measures implemented by the Ministry of Public Health of Qatar also applied, such as temperature checks, social distancing, wearing a mask outside training and matches, and frequent hand hygiene. Club facilities such as showers and recovery areas were closed; players were advised not to use changing rooms and change and shower at home. Indoor activities such as team meetings or gym sessions were limited unless when necessary.

After the quarantine period, teams resumed playing matches, first 1-3 friendly matches, then the final official matches needed to complete the season, with the last match on August 21^st^.

### COVID-19 testing protocol

Before entering and on the day of exiting the quarantine period, players and staff members were submitted to a polymerase chain reaction (PCR) nasopharyngeal swab test (one nasal swab, one pharyngeal swab), performed by a team of specifically trained, experienced nurses using flocked swabs in 3 ml universal transport medium. This was repeated every 3-5 days until the end of the season, scheduled two days before every match. When exiting the hotel quarantine and then approximately every 4 weeks, serum samples were collected for SARS-CoV-2-specific antibody analyses using standard equipment and procedures.

We report test results from two phases, the quarantine phase (entry until exit) and the training and match phase (after quarantine exit until the first test done during the week after the last match on August 21^st^).

### PCR analyses and interpretation

Laboratory analyses were performed centrally in the laboratory facilities of Hamad Medical Corporation in Doha, Qatar. The assays used were pre-validated before routine use in line with the College of American Pathologists accreditation standards. Each platform and reagent combination was analysed for comparative amplification kinetics of each gene target. The following systems were used: (a) EZ1 (QIAGEN, USA) and QIAsymphony (QIAGEN, USA) extraction with thermal cycling using TaqPath™ PCR COVID-19 Combo Kit targeting the N, S and orf1a/b genes (Thermo Fisher Scientific, USA) on ABI 7500 thermal cyclers (Thermo Fisher Scientific, USA); (b) Automated Platforms - the Roche Cobas® 6800 system using the Cobas® SARS-CoV-2 Test targeting the E and orf1a/b genes (Roche, Switzerland) and the Xpert® Xpress SARS-CoV-2 targeting the E and N genes (Cepheid, USA). Reporting followed manufacturer instructions based on the respective cycle threshold (cT) values of each gene-target amplified. Results were reported as positive (cT ≤30), reactive (cT >30 and <40), negative or inconclusive. Reactive was added as a category to identify cases where the likelihood of transmission was deemed low (12–14). The average turnaround between PCR sampling and the result validation and communication was approximately 10 h.

### SARS-CoV-2 antibody analyses

SARS-CoV-2-specific antibodies were measured in serum samples using an electrochemiluminescence immunoassay (Elecsys® Anti-SARS-CoV-2, Roche Diagnostics, Rotkreuz, Switzerland). The assay qualitatively detects SARS-CoV-2 antibodies (IgM and IgG). Tests were interpreted according to the manufacturer’s recommendation, with cut-off indices ≤1 reported as negative and indices >1 as positive. Subjects with positive SARS-CoV-2 antibodies were not required to undergo further PCR testing.

### Results management

Results were communicated automatically to the tested person through the national COVID-19 tracking phone application (mandatory for all inhabitants of Qatar) and, in addition, through their team physicians. If a subject returned a positive or reactive PCR test, he was immediately removed from his team and underwent an isolation protocol implemented by the Ministry of Public Health of Qatar, consisting of 14 days of quarantine for positive cases and 7 days for reactive cases. Upon completion of the quarantine, subjects returned to their normal function within the team and did not have to undergo any further PCR testing (15), but still underwent regular serology testing. Inconclusive samples did not result in any consequences; subjects were just retested on the next scheduled testing date.

Contact tracing was performed by the research team for each case to identify the potential source of infection and other possibly affected subjects.

### COVID-19 symptom reporting

Symptoms were scored for each positive or reactive player using a 26-item symptom score, where symptoms were grouped in three categories: 1) Nose and throat symptoms, 2) Chest and other head/neck symptoms and 3) Whole-body symptoms. The list of specific symptoms is shown in Table 2. Team physicians followed their infected players throughout the isolation period and kept record of their symptoms. Forms were completed on a single occasion by team physicians with the player present once the player was asymptomatic and had returned to play. Players were asked to rate, on average, the severity of each symptom, using a 7-point Likert scale where 1 was “very mild”, 3 “mild”, 5 “moderate” and 7 “severe”, and indicate how many days each symptom lasted. Players could also report “other symptoms”. An overall symptom score was generated as the sum of the scores for each of the three categories.

### Data treatment and ethics

Test results were transferred from the central laboratory to a database adhering to local data safety protocols and analyzed using Tableau Prep Builder 2020 and Tableau Desktop 2020 (Seattle, USA). Population reference data were obtained from the daily COVID-19 numbers officially communicated by the Ministry of Public Health in Qatar throughout the pandemic. We estimated the national incidence of COVID-19 cases in Qatar as the number of PCR-positive or -reactive samples per 100 000 residents per week based on a population of 2.8 million (16). For players, we estimated incidence per 100 000 players per week based on the number of players tested during the study period.

Participating players and staff were not involved in the planning of this research study.

The study was approved by the Institutional Review Board of Aspire Zone Foundation (Application number E202009011).

## Results

### COVID-19 infections in the general population

As shown in Figure 1, the daily number of new PCR-confirmed cases in Qatar was 47 per 100 000 persons on the first day of the study (June 8^th^, 2020), receding gradually during the study period to 17 cases per 100 000 on September 1st. The same was the case for the proportion of positive or reactive PCR samples taken nationally, declining from 29% to 4%. On a weekly basis, the average country-wide rate of infection during the study period was 191 per 100 000 residents per week, with 11.2% positive or reactive among those sampled.

**Figure 1:**
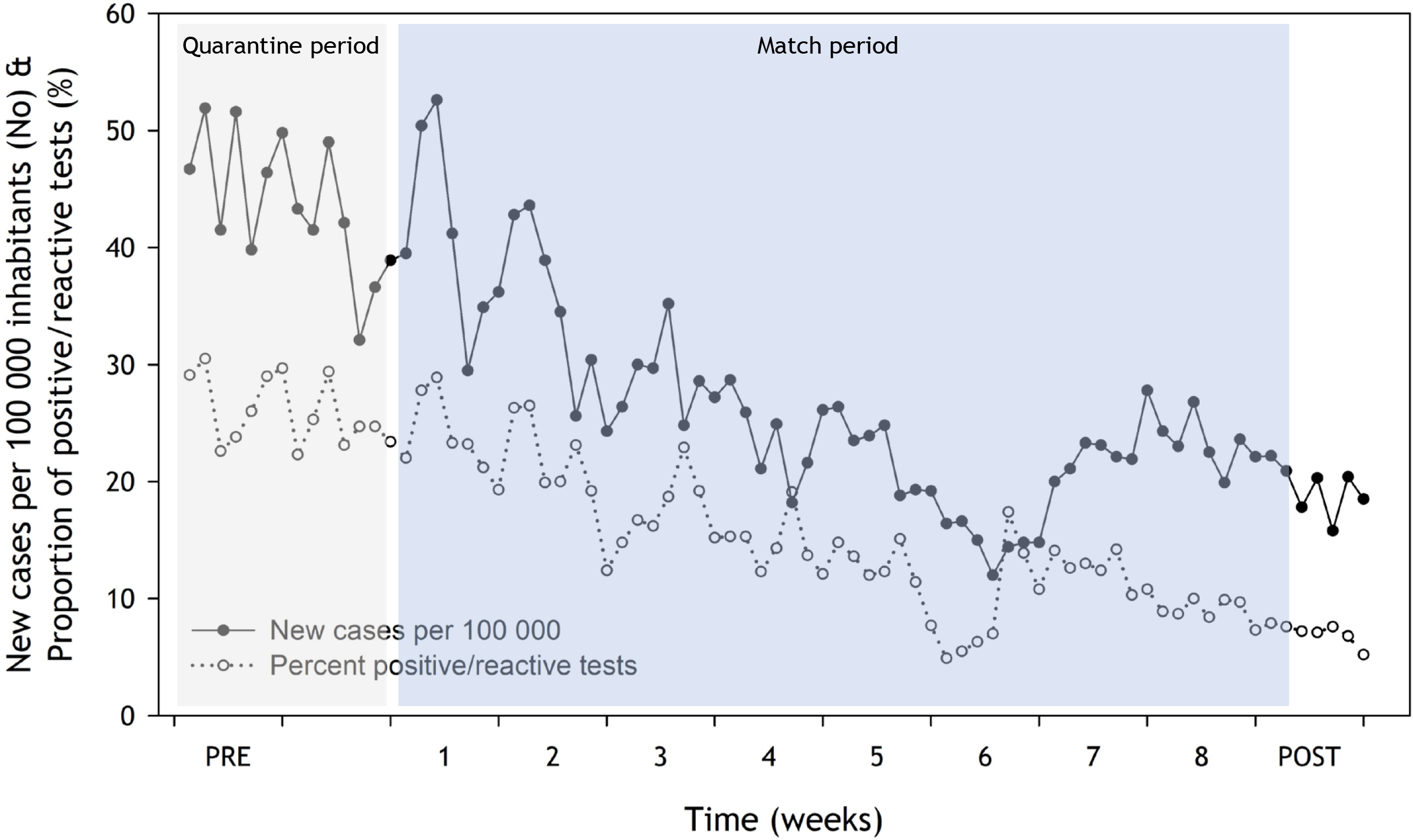
Number of new PCR-confirmed positive or reactive cases (solid symbols & lines, estimated per 100 000 based on a population of 2.8 mill. residents) and percentage positive/reactive of all PCR tests performed in Qatar during the study period (open symbols, dotted lines). The data are based on the report of the analyzing laboratory with 586 905 tests, of which 65 768 were positive or reactive samples (11.2%, corresponding to 191 per 100 000 residents each week on average). PRE denotes the entry test period, starting on June 8^th^, 2020. POST indicates the week after the final match of the season (August 24^th^-September 1^st^). Grey shading denotes the initial hotel quarantine phase; blue shading illustrates the match phase.

### COVID-19 infections among football players and staff

All 12 Division-1 teams and 5 Division-2 teams were included in the study (player age: 26.3±5.3 years (mean ± standard deviation); 54 nationalities). Teams were quarantined for 21±3 days and they resumed inter-team match play 9±6 days after leaving quarantine. Division-1 teams played 102 matches, corresponding to 9±0.5 matches per team; Division-2 teams played 29 matches, 6±2 per team.

In total, 1337 subjects were tested at least once; however, some players and staff joined their team and were gradually included in (or left) the programme during the study period. As shown in Table 1, 757 persons were tested at entry into the quarantine period, with 3.6% positive (N=23) or reactive (N=4) samples, including 7 positive tests among 434 players (1.6%). Subsequent tests prior to and on exit from quarantine, where 719 individuals where tested, revealed an additional 9 positive or reactive samples (1.3%). Of these, 5 were players (1.2%).

**Table 1:** Number of tested persons, positive or reactive tests and percentage of positive/reactive among players and staff at entry into quarantine, exit from quarantine and during the match phase.

|  | Entry into quarantine |  |  | Exit from quarantine |  |  | During match phase |  |  |
| --- | --- | --- | --- | --- | --- | --- | --- | --- | --- |
|  | No. tested | No of Pos./React. | (%) | No. tested | No of Pos./React. | (%) | No. tested | No of Pos./React. | (%) |
| Players | 434 | 7/- | 1.6 | 417 | 1/4 | 1.2 | 549 | 12/12 | 4.4 |
| Medical staff | 82 | 1/1 | 2.4 | 80 | 1/- | 1.3 | 90 | 2/- | 2.2 |
| Technical/coaching staff | 88 | -/3 | 3.4 | 89 | -/1 | 1.1 | 109 | 2/4 | 5.5 |
| Referees | 14 | 3/- | 21.4 | 27 | 1/- | 3.7 | 97 | 1/3 | 4.1 |
| Admin staff | 65 | 7/- | 10.8 | 43 | -/- | 0.0 | 221 | 6/2 | 3.6 |
| Other supporting staff | 74 | 5/- | 6.8 | 63 | -/1 | 1.6 | 101 | 6/- | 5.9 |
| Total | 757 | 23/4 | 3.6 | 719 | 3/6 | 1.3 | 1167 | 28/21 | 4.2 |

During the match phase, between the quarantine exit test and the first test after the final match, 1167 individuals were tested, with 49 (4.2%) positive or reactive tests in total (some players and staff entered the program individually at later stages). Of these, there were 24 positive or reactive tests among the 549 players tested (4.4%). This means that during the study period, excluding the entry screening tests, there were 29 positive or reactive cases among players, corresponding to 457 cases per 100 000 players per week.

Figure 2 displays the timeline for tests performed and test results for the 36 players testing positive or reactive on PCR or serology tests at any time during the study period. Of these, 15 (10 positive, 5 reactive) showed a subsequent positive serology test after their PCR test, thus qualifying as having seroconverted (not all positive or reactive players had a serology test within the timeframe of the study). There were no positive serology tests in players not having a previous positive or reactive PCR test.

**Figure 2:**
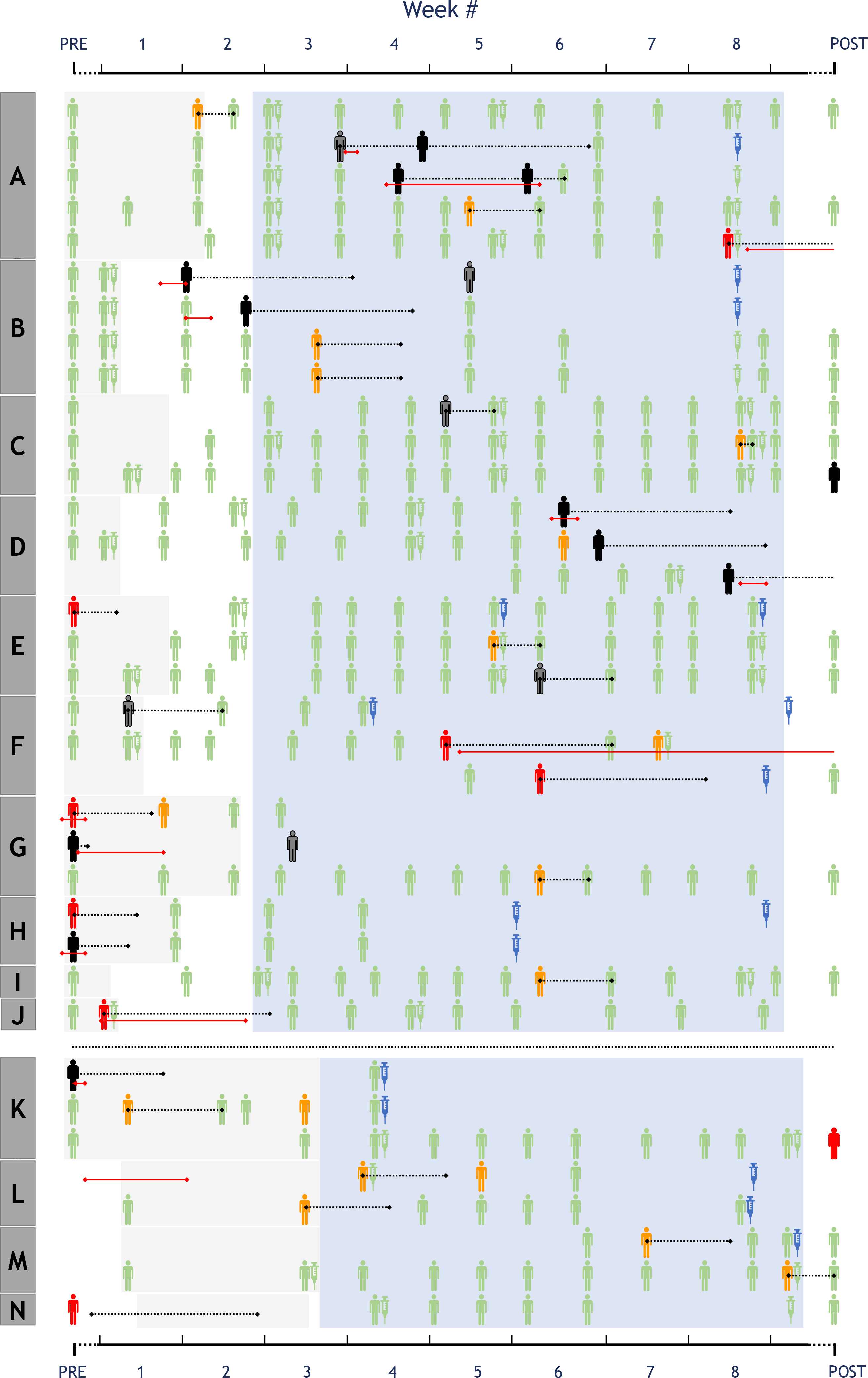
Timeline for tests and test results for the 36 players testing positive or reactive at any time during study period. Every line represents one player, the boxes on the left (A, B, C, etc.) denote their teams. On the horizontal timeline axis, PRE denotes entry tests, performed at the start of each team’s quarantine period, which are shown as shaded grey areas. POST indicates the final test, completed during the week after the final match of the season. Blue shading illustrates the match phase, with Division 1 teams at the top, starting during week 2, and Division 2 teams at the bottom, starting during week 3. Green stick figures depict negative PCR tests, orange reactive and red positive, while black and dark grey figures denote positive and reactive tests traced to sources outside football. Green syringes depict negative serology tests and blue positive (not all positive or reactive players had a follow up serology test by the end of the study period). The horizontal dashed black lines illustrate the individual quarantine period for each player. The red horizontal lines indicate the symptomatic period for players reporting any symptoms.

### Contact tracing

Contact tracing revealed that of the 36 players with positive or reactive PCR tests during the observation period, 20 did not know where or through whom they might have become infected. For 5 players, the infection could be traced to positive family members, and 9 had social interactions (dinner, celebrations) with friends who subsequently returned positive tests. One semi-professional player was infected by a colleague at his workplace. The only confirmed transmission in a football team setting occurred from a physiotherapist (likely infected by his wife who works in the public health sector as a nurse), who subsequently infected one of the players he was treating (other players treated by the same physiotherapist returned negative tests). In another case, two players of the same team were infected at the same time (see Figure 2, team B), likely by each other. As these players had close social contacts (including sharing a car) and none of their teammates were infected, we classified the transmission as “social”. No new infection could be traced to player-to-player transmission during training or matches in this team.

### Symptoms

Of the 36 infected players, 15 reported having symptoms, mainly mild and lasting <1 week (Table 2). None of the 85 individuals (players, staff and referees) testing positive or reactive at any time during the study required hospital admission or medical attention other than limited symptomatic treatment. The association between cT values and symptom scores is shown in Figure 3.

**Table 2:** Number of players reporting symptoms, symptom duration (days) and symptom scores (range 1-7) for the 15 symptomatic players with a positive (11 of 20 players) or reactive test (4 of 16 players).

| Symptom category | Players<br>(n) | No. of<br>days<br>(mean) | Symptom<br>score<br>(mean) |
| --- | --- | --- | --- |
| **Nose and throat symptoms** |  |  |  |
| - Sore throat | 4 | 4 | 3 |
| - Hoarseness | - |  |  |
| - Blocked/plugged nose (nasal congestion) | 2 | 5 | 3 |
| - Runny nose | 3 | 4 | 3 |
| - Sinus pressure | - |  |  |
| - Sneezing | 1 | 4 | 1 |
| - Altered/loss sense of smell | 5 | 14 | 5 |
| - Altered/loss sense of taste | 4 | 12 | 5 |
| **Chest and other head or neck symptoms** |  |  |  |
| - Dry cough | 1 | 5 | 3 |
| - Wet cough i.e. sputum/mucus (coughing up stuff) | - |  |  |
| - Difficulty in breathing (dyspnea) | 1 | 3 | 5 |
| - Fast breathing or shortness of breath | 1 | 7 | 5 |
| - Chest pain | - |  |  |
| - Pressure in the chest | - |  |  |
| - Headache | 9 | 3 | 4 |
| - Conjunctivitis (eye infection) | - |  |  |
| **Whole body symptoms** |  |  |  |
| - Fever | 3 | 2 | 4 |
| - Chills | - |  |  |
| - Excessive tiredness/fatigue | 4 | 5 | 3 |
| - General muscle aches and pains | - |  |  |
| - Skin rash/skin lesions | - |  |  |
| - Abdominal (stomach) pain | - |  |  |
| - Nausea | - |  |  |
| - Vomiting | - |  |  |
| - Diarrhea | 2 | 2 | 3 |
| - Loss of appetite | 1 | 2 | 3 |
| - Other symptoms | - |  |  |

**Figure 3:**
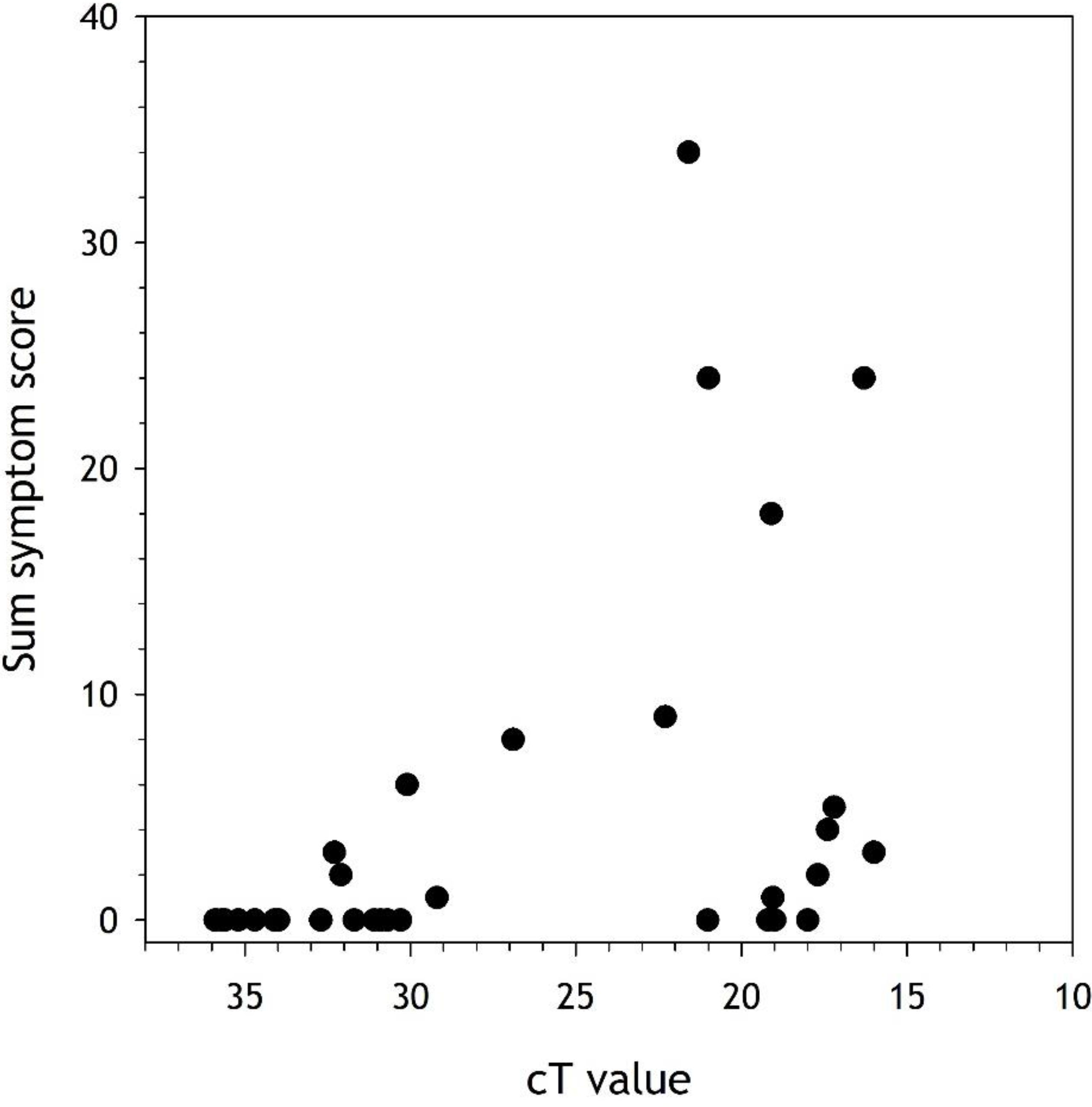
Relationship between the cycle threshold (cT) value and the individual sum of the symptom scores in all positive or reactive players (n=36).

## Discussion

Our study is the first to investigate the risks associated with COVID-19 infection from resuming professional football during the peak of the pandemic in a country with high infection rates. The data show that with the return-to-competition protocol instituted, the season was completed with low risk to player health. The overall infection rate did not exceed what was expected given the viral transmission risk in Qatar at the time, and importantly we did not find any evidence for SARS-CoV-2 transmission during training or matches. Symptoms also were absent or mild in infected players and staff and without hospitalizations or deaths.

### Risk of infections

Whether resuming the football season contributed to an increased risk of infections or not, is difficult to assess with certainty. The country-wide rate of infection in Qatar during the study period was 191 per 100 000 per week, with 11.2% positive or reactive among those sampled. The corresponding figure for players was 457 infections per 100 000 per week (4.4% positive). Since mainly symptomatic individuals were tested in the general population and players were tested regardless of symptoms, this may account for the higher rate of infections found among players during the study period. Fewer than half of the players with positive or reactive tests reported any symptoms.

The German professional football league (Bundesliga) also resumed play at the same time, but with a population infection rate of only 5 per 100 000 per week.(10) It is therefore not surprising that in our collective, 85 of 1337 subjects tested during the entire study period were positive or reactive, whereas in the German study, only 14 of 2506 participants (players and officials) returned positive PCR tests (of which 4 turned out to be remnants of old infections). It is clear there were no mass outbreaks within teams and no matches needed to be cancelled in Qatar in spite of a population infection rate nearly 40 times greater than in Germany.

### Infection pathways

As expected from the asymptomatic course of a large number of infections, we could not determine the source for more than half of our cases. It could be argued that these players were infected during training or matches but testing of the teammates and opposing teams during the period of interest did not reveal any matching infections to suggest player-to-player transmission. The common denominator for the remaining cases is that the infection mainly stemmed from social contacts outside sports, such as friends, family or social events. In such settings, the adherence to preventive measures such as social distancing, wearing a mask, and hand hygiene is often less strict, thus increasing the risk of infection.

In our protocol, we did not find any evidence of transmission during training or matches. It is possible we were unable to trace any infection related to football because of the screening and prevention program itself, and that any positive cases were removed from their team before further infections occurred.

While this was obviously the purpose of the return-to-play protocol, some issues need to be considered. A large number of cases were asymptomatic, and thus not picked up at the daily routine temperature and symptom screening performed by the team physician. Therefore, some infected players likely mixed with their peers before being detected by our testing program. The PCR tests were typically scheduled two days before each match. Based on the known course of infection of SARS-CoV-2, it is possible that subjects returned a negative PCR test at their pre-match screening despite being infected (i.e. incubation period). Two days later, on match day, they could have entered the infectious phase. Therefore, it is likely that some already infected players were not immediately identified by our screening program.

Indeed, one of our cases tested negative two days before his match. He was again tested (outside our program) in the morning of the match at his workplace due to a suspicious case among his colleagues and returned a positive test with a cT value below 20. He was asymptomatic throughout and as the results of the positive workplace test only became available the next day, he played the full match despite being (unknowingly) positive and likely highly infectious. None of his teammates or any member of the opposing team tested positive or reactive within the next two weeks.

This example was representative of many of the positive cases in our players. Owing to the schedule of professional football, with daily team training and 1-2 weekly matches, all of the players testing positive or reactive during the match phase had either trained or played matches within 48 h prior to their test, despite the tight testing scheme and thorough symptom monitoring by team physicians. As can be seen in Figure 3, their cT values and symptom scores ranged widely and some were likely highly infectious at the time of testing. The absence of documented transmission from these infected players to teammates or opposing players supports that the risk of viral transmission during outdoor football is low.

As another example three staff members of one team and one player from another team were infected by a guest during a private dinner. The dinner was held in the evening of one of the testing days, where staff and players had tested negative. They continued to mix with their teams, still tested negative in the next test 4 days later, and were only found positive or reactive 7 days later. None of their teammates and only one staff member was subsequently found to be infected despite completing 5 training sessions and 4 staff meetings over 7 days.

These accounts illustrate that even with a tight testing program there is a risk for having positive and potentially infectious subjects in the population. However, they also support the hypothesis that the infection risk is highest during unprotected exposure in closed spaces, while it is limited when outdoors, even when there is close physical contact between infected and non-infected subjects on the field-of-play. In fact, “contact”, as per infection control criteria, is limited during football matches. During a 90-min match, a player spends only approximately 30-90 seconds in close proximity (<1.5 m) of other players.(8,17) On the other hand, aerosol droplet production is increased with heavy breathing while exercising (9).

There are few descriptions of spreading events related to sports in the literature. Atrubin et al.(18) described an ice hockey match, where 14 out of 22 players and one rink staff member were infected by one player during a single match. Other reports link infections to fitness dance classes and squash facilities (19,20). The main difference between these spreader events and our setting is that they occurred indoors in relatively restricted spaces, whereas our athletes played and trained outdoors. Interestingly, two of the three reports (dance and squash) represent non-contact sports. This supports the hypothesis that respiratory droplets (present in all settings) also represent the main transmission mode during sporting events, but that an outdoor setting with good air circulation attenuates risk.

Thus, when implementing a return-to-play protocol designed to prevent infections, the risk of SARS-CoV-2 transmission during outdoor football training and matches was low.

### COVID-19 disease severity

The infected players showed no or only mild symptoms and none of the participants required hospital admission or medical attention other than limited symptomatic treatment (sore throat, headache etc.). This is in line with the reports of severity in the population of similar age worldwide and in Qatar (21), highlighting the high proportion of asymptomatic or mildly symptomatic cases in young individuals without comorbidities.

Interestingly, even if symptoms were mild in all cases, lower cT values were associated with greater symptom scores (although there were a few exceptions), likely because the cT measured only represents one point in time, while the symptom score retrospectively records the entire course of the infection. Our observation supports previous reports that viral load may be an important determinant for the severity of symptoms of a SARS-CoV-2 infection (12). From a clinical perspective, it suggests to specifically monitor subjects that return low cT values in routine screening tests, as they are more likely to develop more pronounced symptoms. It also highlights that screening interventions based on clinical symptoms only, such as temperature monitoring, might miss a significant number of infections.

### Limitations

This study is limited in that all infections could not be traced to a source. Thus, player to player transmission could not be ruled out with certainty. Still, player transmission during football activities appears unlikely to play a major role in viral spread, given the pattern of infections among the different teams (Figure 2). Clearly, there were no mass outbreaks within teams.

### Conclusion

Our data suggest that professional football played outdoors involving close contact between athletes represents a limited risk for SARS-CoV-2 infection and severe illness when preventive measures are in place, even if the risk of viral transmission in the general population is high. These data may guide organizers of major sports events such as the Olympic Games or continental football championships in their decision-making.

## Data Availability

Data will not be publicly available

## Summary Box

#### Evidence before this study

Professional and leisure team sports were suspended as measures of public health in most countries to prevent the spread of SARS-CoV-2. There is limited information if contact sports played outdoors such as football (soccer) indeed represent an increased risk of viral transmission. The only previous study was conducted in an environment of low viral prevalence.

#### What are the new findings?

We describe viral transmission and the clinical picture of SARS-CoV-2 infections in professional football players and staff in a country which had, at the time of play, among the highest infection rates in the world. Our data shows that with preventive measures in place, the risk of transmission during training and games is low.

Policy makers might consider our results in view of public restrictions to limit viral spread, but also for the assessment of infection risks in the organization of major sports events such as the Olympic Games or professional leagues.

## Acknowledgments

The authors thank the Qatar Stars League for the support of this project.

It would not have been possible without enthusiastic cooperation of the team physicians (Alaaeddine Rahali, Antonio Tramullas, Boudiaf ElHocine, Fawzi Bendimerad, Hafid Mammeri, Hicham Moutaouakkil, Mohammed Nacef, Mokhtar Chaabane, Mondher Barboura, Moumen Jamai Tajdine, Mourad Mokrani, Nebojsa Antic, Noureddine Gharbi, Raouf Rekik, Ovidiu Dragos, Souhail Chebbi, Zied Ellouze) and their medical staff of the participating clubs from the National Sports Medicine Program (NSMP) in implementing the prevention strategy. The tireless efforts of the testing teams directed by Olena Komarcheva were key for this study.

We are grateful to Dr Ian McGuinness and Dr Peter Dzendrowskyj for the stimulating discussions around this project.

We are also indebted to the Laboratory and the Business Intelligence unit of Hamad Medical Corporation for the swift analytical turnaround and results communication.

The 26-item symptom score form was provided by professor Martin Schwellnus, the principal investigator of the AWARE study.

* KC and RB have contributed equally to this work.

## Competing Interests

None.

## Authors and Contributors

YOS, MT, KC and RB have written the first draft of the manuscript, which was checked and edited by KH, IH, AA, PC, AKA, HTB and AJA. PC has performed the analytical measurements, MT, RB, YOS and KC analysed the data. RB, MT, KC, PC and YOS have prepared the figures.

YOS attests that all listed authors meet authorship criteria and that no others meeting the criteria have been omitted.

## Copyright/license for publication

The Corresponding Author has the right to grant on behalf of all authors and does grant on behalf of all authors, a worldwide licence to the Publishers and its licensees in perpetuity, in all forms, formats and media (whether known now or created in the future), to i) publish, reproduce, distribute, display and store the Contribution, ii) translate the Contribution into other languages, create adaptations, reprints, include within collections and create summaries, extracts and/or, abstracts of the Contribution, iii) create any other derivative work(s) based on the Contribution, iv) to exploit all subsidiary rights in the Contribution, v) the inclusion of electronic links from the Contribution to third party material where-ever it may be located; and, vi) licence any third party to do any or all of the above.”

## Transparency statement

YOS (the lead author) affirms that the manuscript is an honest, accurate, and transparent account of the study being reported; that no important aspects of the study have been omitted; and that any discrepancies from the study as originally planned (and, if relevant, registered) have been explained.

## Notes

### Competing Interest Statement

The authors have declared no competing interest.

### Clinical Trial

not registered

### Funding Statement

The study was funded internally

### Summary of Updates

Minor revisions to text, Figure 1 updated

